# The association of covid-19 infection with household food insecurity among Iranian population

**DOI:** 10.1101/2020.12.15.20248221

**Authors:** Naser Kalantari, Neda Ezzeddin, Hassan Eini-Zinab

## Abstract

**Objective:** The aim of this study was to investigate the association of food insecurity score with the status of Covid-19 infection.

**Design:** An online cross-sectional study. Demographic and socio-economic information were collected by questionnaire. Household Food Insecurity Access Scale (HFIAS) was used in assessing household food security status. The analysis data was done by IBM SPSS 22.0, using Chi-square test, ANOVA test and Multinomial logistic regression model.

**Setting:** Iran

**Participants:** 2871 Iranian adults (over 18 years old)

**Results:** findings indicated that men [OR=0.60, CI= (0.41, 0.87), P<0.05], and healthcare personnel [OR=3.66, CI= (1.90, 7.05), P<0.001] were at higher risk for Covid-19. It was also shown that the food insecurity score is significantly higher among infected people compared to not-infected [OR=1.03, CI= (1.00, 1.05), P<0.05]. The comparison between suspected and not-infected individuals only indicated the significant differences in perceived COVID-19 prevention score, which was higher among not-infected people [OR=0.88, CI=(0.84,0.93), P<0.001].

**Conclusion:** Based on the results, in addition to long-term policies to improve food security, policymakers and planners need to plan and implement short-term policies (financial or food assistance) to reduce society vulnerability to the Covid-19.

## Introduction

The novel Covid-19 have had consequences on people life, all over the world ^(1)^.These effects include not only the physical and mental health ^(2)^, but also the livelihoods ^(3)^ and economic status ^(4)^. Many people, specifically self-employed ^(5)^ lost their job or income, which in turn leads to food insecurity ^(6)^. In the study conducted by Kent et.al on Australian, household who lost their income, or had their income reduced at least 25 percent, experienced food insecurity during the pandemic ^(7)^. The pandemic has also disrupted food Supply chain contains production, processing distribution and demand ^(4)^, lead to reduced access to healthy foods ^(8)^ or sometimes increased food prices ^(9)^.

Food insecurity has negative impacts on health ^(10)^, including the function of immune system ^(11,12)^. Given the novelty of covid-19, there is little evidence of an association between the Covid-19 infection and food insecurity. Because the pandemics has been related with increased food insecurity among households from different countries ^(2,13–17)^, this study intends to investigate the association of food insecurity score with the status of Covid-19 infection. The results of the study can be used by policy makers and planners in policy making. It also calls for more attention to food insecurity, especially in Covid-19 infected patients.

## Method

An online cross-sectional study was performed among Iranian society from the whole country, by available sampling method. Participants consist of 2871 Iranian adults (over 18 years old) who invited via social networks such as Telegram Messenger, WhatsApp Messenger and Instagram (from August to September 2020). Before answering the questions, the participants expressed their satisfaction.

Demographic (age, sex and family size), socio-economic (educational level, job and family monthly income) information were collected by questionnaire. Participants were asked to choose their infection status to Covid-19 status from the options: **1**. I have infected (Diagnosis by a physician using PCR test or CT scan tests of the lungs), **2**. I have suspected (Definite infection of someone in close contact; or signs of disease without a diagnostic test in participants) and **3**. None of these conditions (which considered as not-infected). Participants was also ask to score their preventive behavior (Frequent hand washing, wearing a mask, keeping social distance, not attending closed public places, not leaving home except when necessary, etc.) against Covid-19. The range of this score was between 1 (lowest) and 10 (highest), and named as “perceived COVID-19 prevention score” in this study.

Household Food Insecurity Access Scale (HFIAS) is a 9-item questionnaire, used in assessing household food security status, and rated on a Likert scale (0-3). The higher total score of the questionnaire, the greater food insecurity. A standard Persian questionnaire was used in the current study (Cronbach’s alpha=0.85) ^(18)^.

The analysis of data was done by IBM SPSS, Version 22.0 software, using Chi-square test, ANOVA test and Multinomial logistic regression model. Level of less than 0.05 was considered significant. The assessment of data normality was also checked by Kolmogorov–Smirnov test.

## Results

The infection status to Covid-19, among studied people was as follows: infected (N=187, 6.5%), suspected (N=512, 17.8%) and not-infected (N=2172, 75.7%). The mean age and family size of samples were 32.99±8.31 and 3.49±1.29 respectively. The prevalence of women (82.8%) was much higher than men (17.2%), so before the data analyzing, sex weighting was don based on the census data of the Statistics Center of Iran (103 men for every 100 women).

Mean age difference between infected, suspected and not-infected people was assessed via on-way ANOVA test, and it was statistically significant (P<0.05) (table1). The mean family size was also significantly deferent between three groups (P<0.05) (table1). The mean and standard deviation of perceived prevention score was 7.73 ± 1.98, among studied samples and it was higher among not-infected group (P<0.001) (table1). The mean and standard deviation of food insecurity score was 3.60 ± 5.36 and it was higher among infected group compared suspected and not-infected groups (P<0.05) (table1).

**Table 1.** The association of COVID-19 infection with variables

| Quantitative Variables | Infection status to Covid-19 |  |  | P-value<sup>b</sup> |
| --- | --- | --- | --- | --- |
|  | Infected<br>Mean±SD<sup>a</sup><br>( n=187) | Suspected<br>Mean±SD<br>( n=512) | Not-infected<br>Mean±SD<br>( n=2172) |  |
| Age | 32.7±6.6 | 32.1±7.6 | 33.1±8.5 | 0.045 |
| Family size | 3.4±1.3 | 3.6±1.41 | 3.4±1.2 | 0.047 |
| Perceived prevention score | 7.4±2.1 | 7.3±2.0 | 7.8±1.9 | 0.000 |
| Household food insecurity score | 4.5±6.4 | 3.6±5.1 | 3.5±5.3 | 0.034 |
| Qualitative variables | Infected<br>N(%) | Suspected<br>N(%) | Not-infected<br>N(%) | P-value<sup>c</sup> |
| Sex |  |  |  |  |
| Women | 68(4.8) | 248(17.6) | 1090(77.5) | 0.001 |
| Men | 119(8.1) | 264(18) | 1082(73.9) |  |
|  | 187(6.5) | 512(17.8) | 2172(75.7) |  |
| Educational level |  |  |  |  |
| Diploma and lower | 44(7.1) | 110(17.7) | 467(75.2) | 0.883 |
| Associate Degree and Bachelor | 91(6.7) | 241(17.8) | 1021(75.5) |  |
| Masters and higher | 51(5.7) | 162(18) | 685(76.3) |  |
| job status |  |  |  |  |
| student | 11(2.9) | 84(22.3) | 281(74.7) | 0.000 |
| Housewife | 24(4.7) | 100(19.6) | 387(75.7) |  |
| Employee | 58(8.2) | 116(16.3) | 536(75.5) |  |
| Healthcare personnel | 31(13.9) | 39(17.5) | 153(68.6) |  |
| manual worker | 24(11.4) | 37(17.5) | 150(71.1) |  |
| Self-employed | 23(4.3) | 89(16.5) | 427(79.2) |  |
| other | 17(5.6) | 47(15.6) | 238(78.8) |  |
| family income (per month) |  |  |  |  |
| Under 1800000 Rials | 37(7.5) | 83(16.8) | 374(75.7) | 0.808 |
| 18000000-36000000 Rials | 43(6.9) | 102(16.4) | 476(76.7) |  |
| 36000000-54000000 Rials | 36(5.4) | 121(18.1) | 513(76.6) |  |
| 54000000-72000000 Rials | 28(6.9) | 79(19.4) | 300(73.7) |  |
| More than 72000000 Rials | 43(6.3) | 126(18.6) | 509(75.1) |  |
|  | 187(6.5) | 511(17.8) | 2172(75.7) |  |

<sup>a</sup>Standard Deviation
<sup>b</sup>ANOVA
<sup>c</sup> Chi-square test

The educational level of participants included: diploma and lower (21.6%); associate degree and Bachelor (47.1%); and Masters and higher (31.3%). Chi-square test didn’t show any significant association between the educational level and infection status to Covid-19 (P>0.05) (table1). The prevalence of infected, suspected and not-infected individuals by job status, was also examined by Chi-square test. The jobs in order of frequency were: employee (24.7%), self-employed (18.8%), housewife (17.8%), student (13.1%), other jobs (10.5%), healthcare personnel (7.8%) and manual worker (7.3%). The prevalence of infected individuals was higher among healthcare personnel and employees, and self-employed was more prevalent among not-infected people (P<0.001) (table1). There was also a significant difference between groups, by sex. So, the infection status was lower among women (P<0.05) (table1). Monthly family income range was asked from participants and the results were as follows. Under 1800000 Rials (17.2%), 18000000-36000000 Rials (21.6), 36000000-54000000 Rials (23.3), 54000000-72000000 Rials (14.2) and More than 72000000 Rials (23.6). based on the Chi-square test, there wasn’t any difference between the infected, suspected and not-infected participants (P>0.05) (table1).

Based on the results (ANOVA test and Chi-square test), significant predictors were included in multinomial logistic regression models. As shown in table2, the first category of coefficients compares infected versus not-infected; the next category, suspected versus infected. The results indicated that food insecurity score, job status and sex, significantly deference between infected and not-infected people (P<0.05). The comparison between suspected and not-infected individuals only indicated differences in perceived COVID-19 prevention score (P<0.05).

**Table 2.**
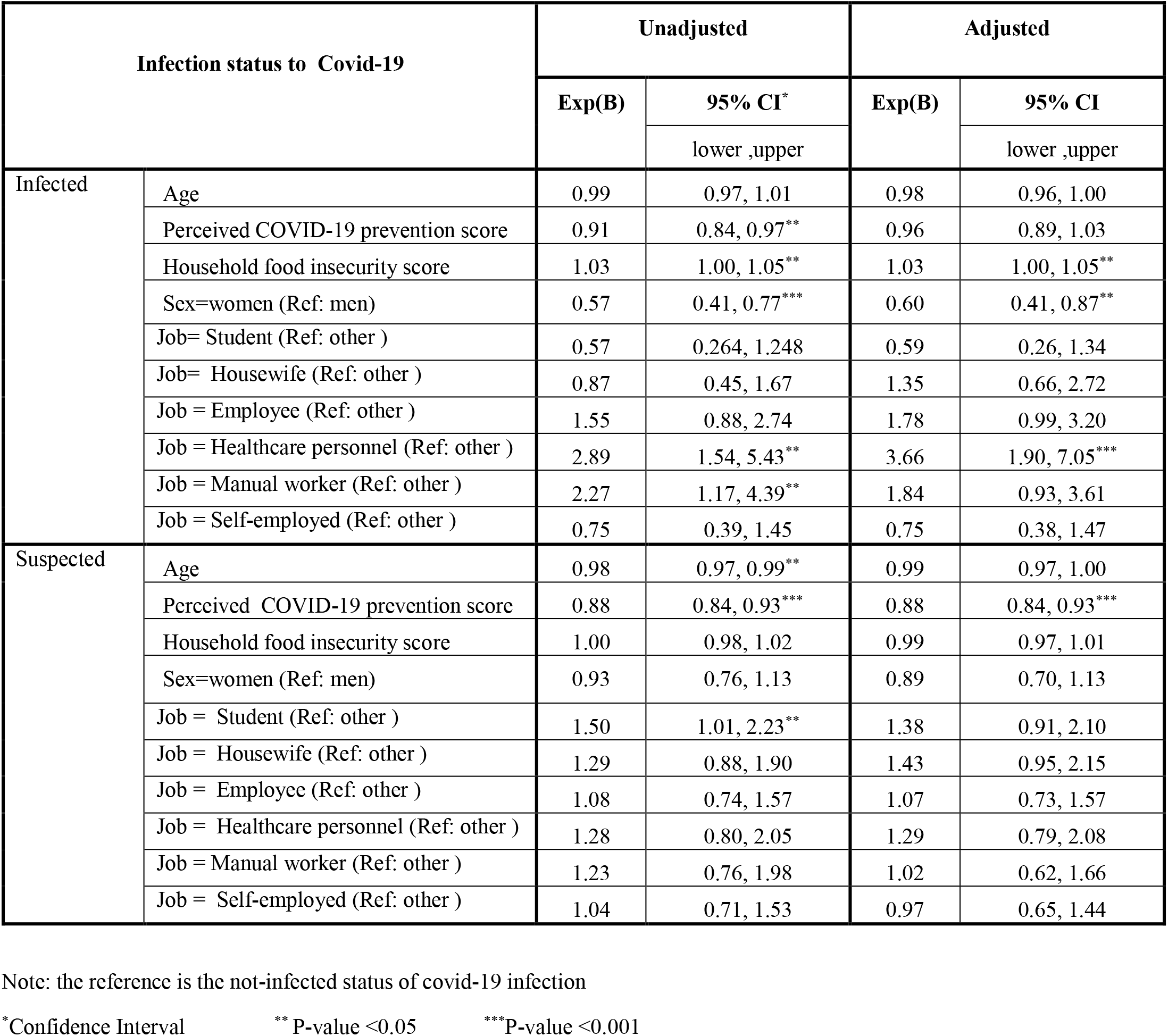
Final Multinomial logistic regression models to examine the association between COVID-19 infection and household food insecurity score

| Infection status to Covid-19 |  | Unadjusted |  | Adjusted |  |
| --- | --- | --- | --- | --- | --- |
|  |  | Exp(B) | 95% CI* | Exp(B) | 95% CI |
|  |  |  | lower ,upper |  | lower ,upper |
| Infected | Age | 0.99 | 0.97, 1.01 | 0.98 | 0.96, 1.00 |
|  | Perceived COVID-19 prevention score | 0.91 | 0.84, 0.97** | 0.96 | 0.89, 1.03 |
|  | Household food insecurity score | 1.03 | 1.00, 1.05** | 1.03 | 1.00, 1.05** |
|  | Sex=women (Ref: men) | 0.57 | 0.41, 0.77*** | 0.60 | 0.41, 0.87** |
|  | Job= Student (Ref: other ) | 0.57 | 0.264, 1.248 | 0.59 | 0.26, 1.34 |
|  | Job= Housewife (Ref: other ) | 0.87 | 0.45, 1.67 | 1.35 | 0.66, 2.72 |
|  | Job = Employee (Ref: other ) | 1.55 | 0.88, 2.74 | 1.78 | 0.99, 3.20 |
|  | Job = Healthcare personnel (Ref: other ) | 2.89 | 1.54, 5.43** | 3.66 | 1.90, 7.05*** |
|  | Job = Manual worker (Ref: other ) | 2.27 | 1.17, 4.39** | 1.84 | 0.93, 3.61 |
|  | Job = Self-employed (Ref: other ) | 0.75 | 0.39, 1.45 | 0.75 | 0.38, 1.47 |
| Suspected | Age | 0.98 | 0.97, 0.99** | 0.99 | 0.97, 1.00 |
|  | Perceived COVID-19 prevention score | 0.88 | 0.84, 0.93*** | 0.88 | 0.84, 0.93*** |
|  | Household food insecurity score | 1.00 | 0.98, 1.02 | 0.99 | 0.97, 1.01 |
|  | Sex=women (Ref: men) | 0.93 | 0.76, 1.13 | 0.89 | 0.70, 1.13 |
|  | Job = Student (Ref: other ) | 1.50 | 1.01, 2.23** | 1.38 | 0.91, 2.10 |
|  | Job = Housewife (Ref: other ) | 1.29 | 0.88, 1.90 | 1.43 | 0.95, 2.15 |
|  | Job = Employee (Ref: other ) | 1.08 | 0.74, 1.57 | 1.07 | 0.73, 1.57 |
|  | Job = Healthcare personnel (Ref: other ) | 1.28 | 0.80, 2.05 | 1.29 | 0.79, 2.08 |
|  | Job = Manual worker (Ref: other ) | 1.23 | 0.76, 1.98 | 1.02 | 0.62, 1.66 |
|  | Job = Self-employed (Ref: other ) | 1.04 | 0.71, 1.53 | 0.97 | 0.65, 1.44 |

Note: the reference is the not-infected status of covid-19 infection
\*Confidence Interval
\*\* P-value <0.05
\*\*\*P-value <0.001

## Discussion

The current study was conducted among Iranian adult population, in order to assess the association of the infection status to Covid-19 and household food insecurity score. Results indicated that men and healthcare personnel were at higher risk for Covid-19. It was also shown that the household food insecurity score was significantly higher among infected people compared to not-infected. This finding is consistent with Escobar et.al study conducted on Californians ^(19)^. The higher infection of people with food insecurity, can be explained from different dimensions. it has been shown that food insecurity can be associated with a weaker immune system response ^(11)^. food insecurity is also associated with greater inflammation in the body ^(20)^. In the study conducted by Kelly et.al, the Ebola deaths was much higher among food insecure individuals ^(12)^. HIV infected people who suffer from food insecurity and Undernutrition were more vulnerable to disease progression ^(21,22)^. There is a similar study on the vulnerability of food insecure children to frequent cold ^(23)^. Leddy et.al showed in a study, that the Antiretroviral Drug function, significantly affected by food security status ^(24)^.

Quality and dietary diversity of food insecure households are lower than food secure ones ^(25,26)^. A healthy diet that provides the micronutrients needed by the body contributes to the better functioning of the immune system ^(27)^. Low-quality diets cause a lack of vital micronutrients for proper body function ^(28)^. Selenium deficiency has been shown to be associated with severe status of Covid-19 infection ^(29)^, through its important role in immune system function ^(30)^. vitamin D deficiency which is related to higher mortality and morbidity due to Covid-19 ^(31–33)^, can be caused by food insecurity ^(34)^ and low family income ^(35)^. The positive effects of vitamin D supplementation have been observed in hospitalized patients with Covid-19 ^(36,37)^.

High calorie, low quality diet in food insecure households, are also associated with overweight and obesity ^(38,39)^. Overweight and obesity predispose people to non-communicable disease ^(40)^. The results of studies performed on hospitalized patients indicated the more vulnerability of obese patient to Covid-19 ^(41–43)^. In the present study, participants’ weight and height information was not collected because the self-reported anthropometric measures accompany with many errors.

Another reason for the higher food insecurity score among infected individuals, may be due to their increased exposure to the virus, because of non-compliance with quarantine. it has been shown that people with lower economic status were less likely to stay at home, during the pandemic ^(44)^. The reason could be their efforts to provide food and basic needs for the family. As seen in the United States, racial and socioeconomic inequalities were evident in infection to Covid-19 ^(45)^. People with lower Scio-economic status, encounter more concerns about employment, income, access to health care and enough food during the pandemic ^(15)^. So, economically vulnerable people may need immediate financial ^(4)^ or food assistance in critical situations, including a pandemic outbreak ^(3)^. However, in the present study, no significant relationship was found between job status and Covid-19 infection, except for health personnel, but further studies are recommended in this area.

In the present study, due to the cross-sectional feature, it is not possible to differentiate between households that suffered from food insecurity before pandemic, households that became food insecure due to pandemic, or post-infection consequences. Therefore, longitudinal studies are recommended to investigate the causal relationship between food insecurity and Covid-19 infection.

The online study has some inevitable limitations, including participation bias. In this study, the participation of young people was higher. The self-reporting form of data gathering, may also have biases. Some people with severe status of COVID-19, may not be able to participate in the study or have not reported their infection, so there may be underreporting in the number of infected people. It is recommended that similar studies, focused on hospitals and health centers, be conducted to examine the association between food insecurity and COVID-19 infection status.

## Conclusion

In the current study, food insecurity score was higher among infected vs. not-infected individuals. This indicates the higher vulnerability of food insecure households against COVID-19. So, in addition to reducing food insecurity and hunger, which is the No. 2 goal of sustainable development ^(46)^, policymakers and planners need to plan and implement short-term policies (financial or food assistance) to reduce society vulnerability to the Covid-19, and increase the resilience to such disaster.

## Data Availability

According to the rules of the research project contract, the researcher is not allowed to share the data, but the data will be available through correspondence with the Vice-Chancellor of Research Affairs

## Acknowledgment

This research grants related to COVID-19, from Vice-Chancellor of Research Affairs, Shahid Beheshti University of Medical Sciences (code: 413, date of approval: 2 August 2020). The authors are thankful from people participated in the study and shared the questionnaire link at Social Networks.

## Ethical Standards Disclosure

This research was approved by Ethics Committee of the National Nutrition and Food Technology Research Institute, Shahid Beheshti University of Medical Sciences (ethical code: IR.SBMU.nnftri.Rec.1399 .028) on 26 July 2020.

